# Rituximab in the treatment of multiple sclerosis in the Hospital District of Southwest Finland

**DOI:** 10.1101/2020.01.15.20017533

**Authors:** Laura Airas, Marjo Nylund, Iina Mannonen, Markus Matilainen, Marcus Sucksdorff, Eero Rissanen

## Abstract

**Background:** There are already numerous B-cell depleting monoclonal anti-CD20 antibodies which have been used to reduce the inflammatory burden associated with multiple sclerosis (MS). We describe here our experience of treating MS-patients with B-cell depleting rituximab.

**Patients and methods:** All MS-patients (n=72) who had received rituximab treatment for at least six months by January 2019 were identified from the patient charts at the Turku University Hospital. Information about MS disease subtype, disease severity, MR-imaging outcomes and B-cell counts were collected from the charts.

**Results:** Rituximab was well received and well tolerated by the patients. There were no serious infusion-related side effects. The most serious adverse event that led to treatment discontinuation was neutropenia. Our study confirms the usability of rituximab treatment for MS in the Finnish health care environment.

**Conclusions:** Off-label rituximab-treatment can be successfully used to reduce MS disease burden for the benefit of MS patients.

## 1. Introduction

B-lymphocytes play an important role in the pathogenesis of MS by providing T-cell help through antigen presentation and cytokine production (Jelcic et al, 2018). In clinical trials B-cell depleting monoclonal antibodies have demonstrated good efficacy and a commendable safety profile in treatment of MS (Hauser et al, 2008; Montalban et al, 2017). Rituximab (MabThera, Roche) is the longest-standing B-cell drug in the market. It is a genetically engineered chimeric monoclonal antibody (mAb) that recognizes CD20 protein on the surface of B lymphocytes. Rituximab binding to the CD20 causes elimination of CD20-expressing B-cells and T-cells (Palanichamy et al, 2014). It has been studied in phase 2 randomized controlled trials in the treatment of RRMS (Hauser et al, 2008) and PPMS (Hawker et al, 2009) (Table 1). In the RRMS study rituximab reduced the amount of contrast enhancing white matter lesions in the brain of MS patients, (Hauser et al, 2008) and the number of relapses decreased more in the rituximab treated patients compared to placebo (Hauser et al, 2008). In the PPMS study, rituximab did not significantly slow down the progression of the disease (Hawker et al, 2009). However, in subgroup analyses, it was shown to slow down the progression of the disease in young (<51 years) PPMS patients and in patients with contrast enhancing lesions (Hawker et al, 2009). The rituximab MS studies did not move on to phase three. Instead, another humanized B-cell depleting mAb, ocrelizumab (Ocrevus, Roche), that was developed based on the rituximab mAb, was investigated in two large phase three studies for MS treatment, and was approved in the European market in the spring of 2018. In addition, a fully human CD20 antibody indicated for the treatment of chronic lymphocytic leukemia, ofatumumab (Arzerra, Novartis), is being studied in two randomized controlled phase 3 trials for treatment of RRMS (Sorensen et al, 2014).

**Table 1.** Summary of studies on B-cell drugs used in the treatment of MS

|  | **Rituximab** | **Ocrelizumab \*\*** | **Ofatumumab** |
| --- | --- | --- | --- |
| **First indication and other indications\*** | -Non-Hodgkin's Lymphoma (NHL)<br>-Chronic lymphocytic leukemia (CLL)<br>- Rheumatoid arthritis<br>-Granulomatous polyangite and microscopic polyangite | Active RRMS<br>Early PPMS | Chronic Lymphocytic Leukemia (CLL) |
| **Phase II studies** | **RRMS**<br>-Hauser et al. HERMES 2008 (Hauser et al, 2008)<br>**PPMS**<br>-Hawker et al. OLYMPUS 2009 (Hawker et al, 2009) | **RRMS**<br>-Kappos et al. 2011 (Kappos et al, 2011)<br>**PPMS \*\*\*** | **RRMS**<br>-Sorensen et al. 2014 (Sorensen et al, 2014) |
| **Phase III studies** | - | **RRMS**<br>Hauser et al. 2017<br>OPERA I & II 2016 (Hauser et al, 2017)<br>**PPMS**<br>-Montalban et al. 2017<br>ORATORIO 2017 (Montalban et al, 2017) | **RRMS**<br>Hauser et al. 2019<br>ASCLEPIOS I and II 2019 (Hauser et al, 2019) |
| **EMA Approval** | - | Jan 8, 2018 | - |

RRMS = relapsing remitting MS, PPMS = Primary Progressive MS, EMA = European Medicines Agency
\* Rituximab was originally developed for the treatment of non-Hodgkin's lymphoma in 1997, but its indications have expanded. In addition, it is also used as an off-label treatment for many other autoimmune diseases (36-38). The exact total number of rituximab-treated patients is unknown, but there are already over 300,000 rituximab-treated rheumatoid arthritis patients worldwide.
\*\* The excellent efficacy of ocrelizumab in the treatment of RRMS was demonstrated in two large phase III trials (OPERA I and II) (35). In an extensive phase III PPMS study, ocrelizumab was shown to be the first MS treatment that reduces progression of disability in patients with progressive MS (ORATORIO) (6).
\*\*\* Phase 2 PPMS trial was not performed with ocrelizumab, but phase 3 studies were directly performed based on the results of the rituximab phase 2 study.

Due to the high efficacy, safety and convenient administration (one infusion every 6 months) of B-cell depleting medications, and the relatively low cost of rituximab compared to other MS medications, the B-cell drugs were introduced in treatment of MS already before ocrelizumab was officially authorized. Due to this, there is abundant published real-world research data about rituximab experience in MS treatment (Alcala et al, 2018; Alldredge et al, 2018; Barra et al, 2016; D’Amico et al, 2018; Rommer et al, 2016). In Sweden, off-label use of rituximab in the treatment of MS patients has become common and the Swedish experience has been reported in large retrospective registry studies (Alping et al, 2016; Salzer et al, 2016b). Rituximab is given as an intravenous infusion every six months, with the most common single doses for MS being 500 mg or 1000 mg. The Swedish MS Association has guidelines for the practical implementation of rituximab medication (http://www.mssallskapet.se) and we have also applied these guidelines in the Hospital District of Southwest Finland (Table 2).

**Table 2.** Practical implementation of rituximab treatment in Turku University Hospital

|  |  |
| --- | --- |
| **Prior treatment initiation** | – Check the patient's vaccination status<br>– If necessary instruct vaccination and wait at least 4 weeks before rituximab infusion.<br>– Safety blood samples prior to treatment<br>• Blood white cell counts with differentials, CRP, creatine, ALT, serum anti-VZV antibodies, lymphocyte subtyping for CD4, CD8 and CD19 cells, NK cells and monocytes, Quantiferon assay, serum anti-JCV antibodies, serum anti-hepatitis B antibodies, plasma IgG-level<br>• Immediately prior to infusion, acute infections are ruled out, if necessary by laboratory testing |
| **Adjuvant therapy for the infusion** | – Before rituximab infusion Solu-Medrol 250 mg in 100 ml of NaCl 0.9% is given i.v., so that the infusion is finished ½ hour before the initiation of rituximab infusion.<br>– 1 g paracetamol and 10 mg cetirizin are given orally approximately ½ hour before the rituximab infusion.<br>– Blood pressure monitoring is performed every ½ hour, prior to the start of the infusion and always prior to dose increase (as per SPC guidelines) |
| **Dosage** | – The first dose is 1000 mg, then 500 mg every 6 months for 3 years, or at the discretion of the treating neurologist<br>– Rituximab infusion is administered via a drip counter. |
| **Follow-up blood tests** | – 3 months after infusion:<br>• Blood cell counts with differentials, and lymphocyte subtyping<br>– 6 months after infusion<br>• Blood cell counts with differentials, lymphocyte subtyping and plasma immunoglobulin levels |

## 2. Materials and methods

This is a single center retrospective observational study on the usability of rituximab in the treatment of MS in Finland. Rituximab therapy was offered in the area of the Hospital District of Southwest Finland during years 2014–2019 for MS patients for whom other MS medications had failed to achieve a desired treatment effect, or for whom no other suitable medication was available. All treated patients were aware that the drug had no official indication for the treatment of MS. MS patients who had received at least two doses of rituximab by January 2019 were included in the study. Information about medication, severity of MS, relapses, infections and adverse reactions during the treatment, infusion reactions, MRI results and laboratory measurements were collected from the medical records of the Hospital District of Southwest Finland. CD19-positive B-cells were assayed using flow cytometry in local hospital laboratory (Tykslab). Expanded Disability Status Scale (EDSS) scores for disability assessment were collected from medical records. The number of relapses was obtained from the medical records two years before and one year after rituximab initiation. Brain MRIs were evaluated for the number of contrast enhancing lesions. The study was approved by the Hospital District of Southwest Finland. In the statistical evaluation of relapses, EDSS and MRI changes, different time points were compared by the MS subtype using the Wilcoxon signed-rank test, adjusted with Holm method. A p-value <0.05 was considered statistically significant.

## 3. Results

A total of 72 MS patients met the criteria of the study. Of these, 31 had RRMS, 16 had PPMS and 25 had SPMS, and the study covered a total of 127.7 patient years. Table 3 provides a description of the patient characteristics. 73.6 % of patients were under 51 years of age. 76.4 % of patients (n = 55) had available MRI results just prior to initiation of medication. Approximately one-fourth (26.9 %; n = 18) of them had contrast-enhancing lesions. Table 2 describes the dosing and monitoring instructions for rituximab medication in the Hospital District of Southwest Finland. The drug was administered as an intravenous infusion in doses of 500 mg or 1000 mg according to the instruction of the treating neurologist, and the interval between the first and second infusions was on average 7 months. 45.8 % of all patients had treatment initiated with the 1000 mg dose. Among the RRMS patients, a second infusion was given after 6 months (median; range 6-9). Subsequent doses and timing of administration was decided by the treating neurologist. Dose intervals ranged from 5 months to 21 months, with a median dose interval of 7 months. There was no significant difference between the MS disease subgroups in the dose sizes or dose intervals. The study patients received rituximab treatment for 1.7 years (median; range 0.6 - 3.8) and 85 % of patients received rituximab treatment for at least 1 year.

**Table 3.** Demographic description

|  |  | **RRMS**<br>n = 31 | **PPMS**<br>n = 16 | **SPMS**<br>n = 25 | **All**<br>n = 72 |
| --- | --- | --- | --- | --- | --- |
| Female, n (%) |  | 22 (71.0) | 7 (43.8) | 14 (56.0) | 43 (59.7) |
| Age at first symptoms, y | mean (sd) | 31.9 (11.0) | 39.1 (11.3) | 30.0 (10.7) | 33.0 (11.3) |
|  | range | 15.1-54.1 | 22.2-59.9 | 17.0-59.1 | 15.1-59.9 |
| Age at dg, y | mean (sd) | 34.7 (12.1) | 41.8 (12.1) | 31.5 (11.4) | 35.2 (12.3) |
|  | range | 15.4-57.7 | 22.9-61.9 | 17.0-66.3 | 15.4-66.3 |
| Age RTX, y | mean (sd) | 39.8 (11.9) | 44.7 (11.8) | 47.8 (9.5) | 43.7 (11.5) |
|  | range | 17.6-59.0 | 24.2-62.5 | 27.0-66.5 | 17.6-66.5 |
| Disease duration, y | mean (sd) | 7.8 (6.6) | 5.6 (5.3) | 18.2 (8.1) | 11.0 (8.7) |
|  | range | 0.3-23.2 | 0.6-21.2 | 7.4-39.3 | 0.3-39.3 |
| SPMS duration, y | mean (sd) | - | - | 6.0 (4.0) | - |
|  | range |  |  | 0.23-16.8 |  |
| EDSS at baseline* | md (range) | 3 (1-7) | 5 (2-7) | 6 (3-8) | 4.5 (1-8) |
| No of RTX infusions | md (range) | 3 (2-6) | 3.5 (2-6) | 3 (2-6) | 3 (2-6) |
| First RTX dosage 500 mg | n (%) | 17 (54.8) | 7 (43.8) | 15 (60.0) | 39 (54.2) |
| ARR 2y before RTX | mean (sd) | 0.73 (0.56) | 0.03 (0.13) | 0.29 (0.39) | 0.42 (0.52) |
|  | range | 0-2 | 0-0.5 | 0-1 | 0-2 |
| ARR 1y during RTX | mean (sd) | 0.13 (0.34) | 0.13 (0.5) | 0.08 (0.28) | 0.11 (0.36) |
|  | range | 0-1 | 0-2 | 0-1 | 0-2 |
| JCV-positive | n (%) | 22 (71.0) | 10 (83.3) | 12 (57.1) | 44 (68.8) |
| Treatment naïve | n (%) | 7 (22.6) | 12 (75.0) | 3 (12.0) | 22 (30.6) |
| Previous treatment | n (%) |  |  |  |  |
| Natalizumab |  | 9 (29.0) | 2 (12.5) | 3 (12.0) | 14 (19.4) |
| Beta-interferon |  | 2 (6.5) | 0 (0.0) | 1 (4.0) | 3 (4.2) |
| Glatiramer acetate |  | 5 (16.1) | 1 (6.3) | 2 (8.0) | 8 (11.1) |
| Fingolimod |  | 6 (19.4) | 0 (0.0) | 10 (40.0) | 16 (22.2) |
| Other |  | 2 (6.5) | 1 (6.3) | 6 (24.0) | 9 (12.5) |
| Follow-up time, y | mean (sd) | 1.5 (0.7) | 2.3 (0.9) | 1.9 (0.8) | 1.9 (0.8) |
|  | range | 0.6-3.8 | 0.8-4.2 | 0.7-3.4 | 0.6-4.2 |
| RTX discontinuation during follow-up | n (%) | 1 (3.2) | 4 (25.0) | 7 (28.0) | 12 (16.7) |

sd = standard deviation, md = median, RTX = rituximab, RRMS = relapsing remitting MS, PPMS = primary progressive MS, SPMS = secondary progressive MS, EDSS = Expanded Disability Status Scale, JCV = John Cunningham virus
\*EDSS was available for 64 patients

A total of 12 patients discontinued medication during the follow-up. 11 patients who discontinued treatment had progressive MS (5 PPMS and 6 SPMS patients). The reason for discontinuation was either patient’s disappointment with the drug efficacy (n = 10), or a drug-related adverse event (n = 2). During the study, rituximab was administered a total of 233 times, of which 48 was a 1000 mg dose.

### 3.1. B-cell levels and infusion frequency

The effect of rituximab treatment on blood cells was monitored by regular laboratory tests. The purpose of the laboratory follow-up was to exclude the possibility of a neutropenic adverse effect and to evaluate the effect of the treatment on B-cell counts. B-cells are known to deplete rapidly after the first infusion of rituximab (Hauser et al, 2008), and a rapid and sustained reduction of B-cells was also seen in this study (Figure 1D). B-cell levels influenced the treatment decisions of the treating neurologist. In patients with very low B-cells (< 10 x 10^6^/l) for longer periods of time (> 6 months), the infusion interval was often prolonged.

**Figure 1.**
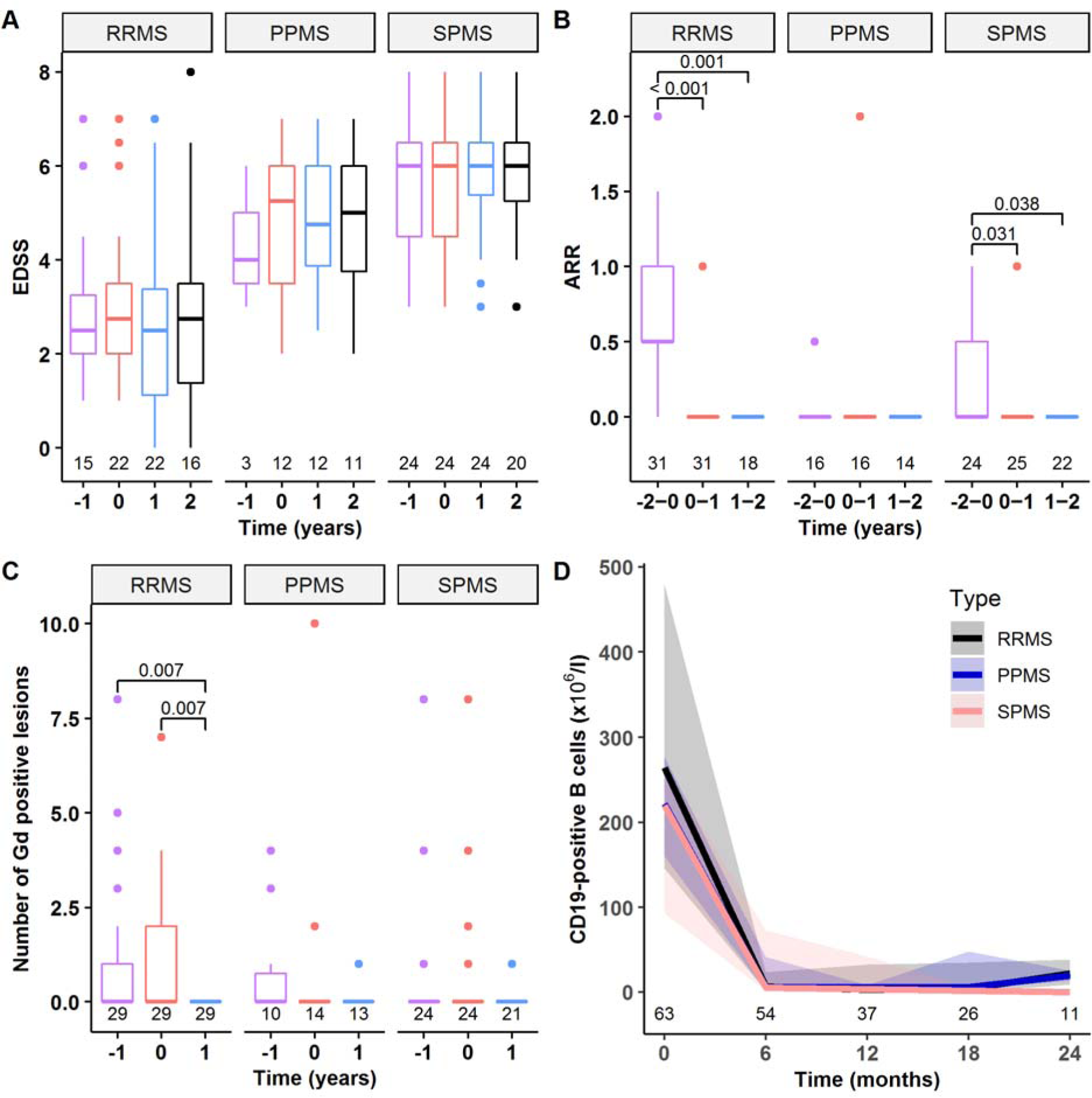
EDSS, relapses, contrast enhancing lesions and B-cell counts before and during rituximab treatment. **A**. EDSS during rituximab treatment (1 and 2 years after initiation), at initiation of treatment (0) and 1 year prior initiation of treatment (−1 years). **B**. Mean number of relapses per year 2 years before and after initiation of treatment. **C**. The number of contrast-enhancing lesions in MRI taken approximately one year before initiation of treatment (−1), just prior to initiation of treatment (0), and approximately one year after initiation of treatment (1). **D**. Median CD19 positive B-cell counts (with 1^st^ and 3^rd^ quartiles) just prior to treatment and every six months after initiation of treatment. The median number of CD19-positive B-cells 6 months after the first infusion was 6 x10^6^/l (quartiles 1 and 45); normal range 90-660 x10^6^/l (Tykslab). In the graphs A-C, the p-values represent statistically significant differences between the time points (Wilcoxon signed-rank test, adjusted with Holm method). RRMS = Relapsing remitting MS, PPMS = Primary Progressive MS, SPMS = Secondary Progressive MS, EDSS = Expanded Disability Status Scale, Gd+ = Gadolinium enhancing lesion. The number of patients is shown in graphs.

### 3.2. Patients’ functional performance during rituximab treatment

Patients’ functional performance remained stable during rituximab treatment in all MS groups as measured by EDSS (Figure 1A). No statistically significant changes were observed in EDSS during treatment when compared to pre-treatment values (Figure 1A). 31 % of the RRMS patients had reduced disability which was demonstrated as a decrease in EDSS, 13 % had an increase in EDSS, and in 56 % of the RRMS patients the EDSS score remained unchanged from baseline to the end of follow up. Among the PPMS patients 18 % had reduced disability, 36 % had increased disability and 45 % had stable EDSS during the study. Among SPMS patients, the corresponding figures were 20 %, 35 % and 45 %.

### 3.3. Number of relapses during rituximab treatment

The annualized relapse rate (ARR) was obtained from the medical records for the two years before the treatment initiation and for the first and second years of rituximab treatment (Figure 1B). In both RRMS and SPMS patient groups, there was a statistically significant reduction in relapses following the initiation of rituximab treatment (Figure 1B).

### 3.4. Contrast enhancing lesions in MRI

Approximately one-fourth (26.9 %; n = 18) of subjects had contrast enhancing lesions at baseline MRI. The reduction in enhancing lesions after treatment initiation was statistically significant among RRMS patients (Figure 1C).

### 3.5. Side effects associated with rituximab

41.7% of patients (n = 30) experienced minor infusion-related reactions during rituximab administration. The reactions were often mild flu-like symptoms, dizziness, or pruritus, and they occurred most frequently with the first infusion. In individual cases (n = 4), infusion-related side effects were more pronounced. One patient experienced confusion and spasticity, another experienced dyspnoea and two experienced tachycardia.

One SPMS patient developed severe neutropenia and an unspecified bacterial infection after fourth rituximab infusion (neutrophil count 0.00 x 10^9^/ l). The infection was successfully treated with antibiotics and neutropenia recovered after treatment with neutrophil growth factor therapy. Another patient at our hospital developed neutropenia immediately after the first infusion of rituximab, which led to discontinuation of the drug (Rissanen et al, 2017).

The most common infections during rituximab therapy were lower urinary tract infections (n = 12) and various respiratory infections (upper respiratory infections (n = 6) and pneumonia (n = 3)). In addition, one subject had influenza, two subjects developed dental infection and one patient developed an infectious abscess in the jaw area (Table 4).

**Table 4.** Infections during rituximab treatment.

|  | **RRMS**<br>n = 31 | **PPMS**<br>n = 16 | **SPMS**<br>n = 25 | **All**<br>n = 72 |
| --- | --- | --- | --- | --- |
| Lower urinary tract infection, n (%) | 3 (10 %) | 4 (25 %) | 5 (20 %) | 12 (17 %) |
| Pneumonia, n (%) | 2 (6 %) | 0 | 1 (4 %) | 3 (4 %) |
| Upper respiratory infection, n (%) | 4 (13 %) | 1 (6 %) | 1 (4 %) | 6 (8 %) |
| Other, n (%) | 5 (16 %) | 4 (25 %) | 3 (12%) | 12 (16 %) |
| Other infections included 2 influenza, 1 abscess, 2 dental infections, 1 pyelonephritis, 1 herpes, 2 asymptomatic sinusitis and 3 undetermined infection. RRMS = relapsing remitting MS, PPMS = primary progressive MS, SPMS = secondary progressive MS |  |  |  |  |

## 4. Discussion

Off-label use of rituximab in the treatment of MS is widespread throughout the world (Alcala et al, 2018; Alldredge et al, 2018; Barra et al, 2016; Berenguer-Ruiz et al, 2016; D’Amico et al, 2018; de Flon et al, 2016; de Flon et al, 2017; Memon et al, 2018; Nielsen et al, 2012; Rommer et al, 2016; Salzer et al, 2016a; Salzer et al, 2016b; Scotti et al, 2018; Topping et al, 2016), and our retrospective study indicates that rituximab is also a usable treatment option among Finnish MS patients. Most of our patients were satisfied with the effectiveness and the easy administration of the treatment, and were generally willing to initiate this treatment despite its off-label status as MS treatment. This study includes patients from a time when it was not yet possible to use ocrelizumab for treatment of MS. Today, however, the excellent phase 3 ocrelizumab study results, its good safety profile and the positive real-world experience gained so far, support the usability of ocrelizumab as a similarly convenient, effective and safe B-cell therapy for MS.

There are few guidelines and regulations on the use of off-label medicines in Finland (Hermanson, 2008). According to the regulation of the Ministry of Social Affairs and Health (726/2003), the physician must pay particular attention to the efficacy, safety and price of the selected medication, but the regulations do not oblige the physician to adhere to the official label (Hermanson, 2008). Recent current care guidelines of MS in Finland suggests that rituximab may be considered for the treatment of MS in individual cases (Finnish Current Care Guidelines, 2019). It is good practice to inform the patient about the off-label nature of the treatment, to state this in the patient charts, and to make the final treatment decisions together with the patient. The individual risk-benefit ratio of the medication is carefully evaluated, and affects the choice of medication, and health economic aspects can also be taken into account when choosing the treatment. On the other hand, in many other European countries local laws and regulations, or lack of reimbursement entirely prevent the use of off-label medication (Marjolein et al, 2019). Interestingly, 90% of all Finnish rituximab-treated MS-patients are in the area of Hospital District of Southwest Finland. Ocrelizumab-treatment on the other hand is more evenly distributed with approximately 3 % of all Finnish MS-patients being treated with this medication.

Rituximab-treatment was relatively well tolerated among our patient cohort. Delayed neutropenia was the most serious side effect. Deep neutropenia can be treated with neutrophil growth factor therapy, but if untreated, it can lead to severe infection requiring hospitalization (Breuer et al, 2014). Therefore, we recommend a white blood cell count every 3 months. In addition, our patients are instructed to seek laboratory blood tests if symptoms of infection occur in order to rule out possible neutropenia. B-cell depletion will inevitably lead to some degree of immunosuppression, but serious opportunistic infections are rare with B-cell therapy. No unconfounded cases of PML have been reported in MS patients in association with rituximab-treatment. In rheumatoid arthritis cohort, the PML-risk is estimated to be 1: 25000, but here rituximab treatment is often combined with other immunosuppressive therapy, which may increase the risk of PML. In the Swedish cohort, pneumonia occurred in 14% of patients receiving rituximab (Clifford et al, 2011). Longer-term treatment with B-cell depleting drugs may lead to hypogammaglobulinemia, which also predisposes to infections. If a sustained decrease in plasma IgG antibody levels is observed, immunoglobulin replacement therapy may be given intravenously. B-cell depleting drugs attenuate vaccine responses (Arad et al, 2011; Hua et al, 2014), and we recommend vaccination against pneumococcus and updating of other vaccines before starting B-cell treatment. In the initial rituximab study, RRMS patients were dosed twice with 1000 mg with a two-week interval (Hauser et al, 2008). In later real-world studies, also lower doses of rituximab have proven efficient (Alping et al, 2016; Svenningsson et al, 2015). Monitoring of B-cell levels may help in the assessment of drug response, and if the number of CD19-positive cells has increased markedly 6 months after the last infusion, a dose increase from 500 mg to 1000 mg may be necessary. On the other hand, if the patient has an increased incidence of infections during rituximab treatment, it may be warranted to prolong the infusion interval, especially if B-cell levels are consistently low. Longer dosing intervals may reduce susceptibility to infections and reduce the risk of hypogammaglobulinemia.

## 5. Conclusions

The pathogenesis of MS is a continuum of inflammation and neurodegeneration. Relapses are a clinical manifestation of acute central nervous system inflammation, and steady symptom progression is associated with more chronic brain pathology. Inflammation and neurodegeneration are closely related, and it is likely that the benefits of B-cell therapy are primarily related to the prevention of inflammation even in the progressive forms of the disease. There are small differences between B-cell drugs, but they all work by binding to the CD20 molecule and by causing depletion of B-cells. Theoretically, B-cell depleting drugs are all effective and well-tolerated in the treatment of MS, but the level of scientific evidence on the efficacy varies between the drugs (Table 1). Direct head-to-head trials between the drugs have not been performed. MS-treating doctors in certain European countries are free to choose the treatment for their patients from a wider variety of drugs, including off-label treatments, whereas other colleagues are more limited due to local laws and regulations.

MS is a life-long disease that requires long-term treatment, and time will tell the true efficacy of the B-cell drugs in long-term care, and uncover any potential hidden drug-related drawbacks.

## Data Availability

The anonymized raw data not published within the article will be shared over the next 3 years upon request from a qualified investigator.

## References

Alcala, C., Gascon, F., Perez-Miralles, F., Gil-Perotin, S., Navarre, A., Bosca, I., Coret, F. & Casanova, B. (2018) Efficacy and safety of rituximab in relapsing and progressive multiple sclerosis: a hospital-based study. J Neurol, 265(7), 1690–1697.

Alldredge, B., Jordan, A., Imitola, J. & Racke, M. K. (2018) Safety and Efficacy of Rituximab: Experience of a Single Multiple Sclerosis Center. Clin Neuropharmacol, 41(2), 56–59.

Alping, P., Frisell, T., Novakova, L., Islam-Jakobsson, P., Salzer, J., Björck, A., Axelsson, M., Malmeström, C., Fink, K., Lycke, J., Svenningsson, A. & Piehl, F. (2016) Rituximab versus fingolimod after natalizumab in multiple sclerosis patients. Ann Neurol, 79(6), 950–8.

Arad, U., Tzadok, S., Amir, S., Mandelboim, M., Mendelson, E., Wigler, I., Sarbagil-Maman, H., Paran, D., Caspi, D. & Elkayam, O. (2011) The cellular immune response to influenza vaccination is preserved in rheumatoid arthritis patients treated with rituximab. Vaccine, 29(8), 1643–8.

Barra, M. E., Soni, D., Vo, K. H., Chitnis, T. & Stankiewicz, J. M. (2016) Experience with long-term rituximab use in a multiple sclerosis clinic. Mult Scler J Exp Transl Clin, 2, 2055217316672100.

Berenguer-Ruiz, L., Sempere, A. P., Gimenez-Martinez, J., Gabaldon-Torres, L., Tahoces, L., Sanchez-Perez, R. & Diaz-Marin, C. (2016) Rescue Therapy Using Rituximab for Multiple Sclerosis. Clin Neuropharmacol, 39(4), 178–81.

Breuer, G. S., Ehrenfeld, M., Rosner, I., Balbir-Gurman, A., Zisman, D., Oren, S. & Paran, D. (2014) Late-onset neutropenia following rituximab treatment for rheumatologic conditions. Clin Rheumatol, 33(9), 1337–40.

Clifford, D. B., Ances, B., Costello, C., Rosen-Schmidt, S., Andersson, M., Parks, D., Perry, A., Yerra, R., Schmidt, R., Alvarez, E. & Tyler, K. L. (2011) Rituximab-associated progressive multifocal leukoencephalopathy in rheumatoid arthritis. Arch Neurol, 68(9), 1156–64.

D’Amico, E., Zanghi, A., Chisari, C. G., Fermo, S. L., Toscano, S., Arena, S., Patti, F. & Zappia, M. (2018) Effectiveness and safety of Rituximab in demyelinating diseases spectrum: An Italian experience. Mult Scler Relat Disord, 27, 324–326.

de Flon, P., Gunnarsson, M., Laurell, K., Soderstrom, L., Birgander, R., Lindqvist, T., Krauss, W., Dring, A., Bergman, J., Sundstrom, P. & Svenningsson, A. (2016) Reduced inflammation in relapsing-remitting multiple sclerosis after therapy switch to rituximab. Neurology, 87(2), 141–7.

de Flon, P., Laurell, K., Soderstrom, L., Gunnarsson, M. & Svenningsson, A. (2017) Improved treatment satisfaction after switching therapy to rituximab in relapsing-remitting MS. Mult Scler, 23(9), 1249–1257.

Hauser, S. L. (Oral presentation. ECTRIMS 2019) Efficacy and safety of ofatumumab versus teriflunomide in relapsing multiple sclerosis: results of the phase 3 ASCLEPIOS I and II trials.

Hauser, S. L., Bar-Or, A., Comi, G., Giovannoni, G., Hartung, H. P., Hemmer, B., Lublin, F., Montalban, X., Rammohan, K. W., Selmaj, K., Traboulsee, A., Wolinsky, J. S., Arnold, D. L., Klingelschmitt, G., Masterman, D., Fontoura, P., Belachew, S., Chin, P., Mairon, N., Garren, H., Kappos, L. & Investigators, O. I. a. O. I. C. (2017) Ocrelizumab versus Interferon Beta-1a in Relapsing Multiple Sclerosis. N Engl J Med, 376(3), 221–234.

Hauser, S. L., Waubant, E., Arnold, D. L., Vollmer, T., Antel, J., Fox, R. J., Bar-Or, A., Panzara, M., Sarkar, N., Agarwal, S., Langer-Gould, A., Smith, C. H. & Group, H. T. (2008) B-cell depletion with rituximab in relapsing-remitting multiple sclerosis. N Engl J Med, 358(7), 676–88.

Hawker, K., O’Connor, P., Freedman, M. S., Calabresi, P. A., Antel, J., Simon, J., Hauser, S., Waubant, E., Vollmer, T., Panitch, H., Zhang, J., Chin, P. & Smith, C. H. (2009) Rituximab in patients with primary progressive multiple sclerosis: results of a randomized double-blind placebo-controlled multicenter trial. Ann Neurol, 66(4), 460–71.

Hermanson, T. (2008) Saako lääkettä määrätä myyntiluvasta poiketen? Lääketieteellinen Aikakauskirja Duodecim, 124(24), 2777–8.

Hua, C., Barnetche, T., Combe, B. & Morel, J. (2014) Effect of methotrexate, anti-tumor necrosis factor alpha, and rituximab on the immune response to influenza and pneumococcal vaccines in patients with rheumatoid arthritis: a systematic review and meta-analysis. Arthritis Care Res (Hoboken), 66(7), 1016–26.

Jelcic, I., Al Nimer, F., Wang, J., Lentsch, V., Planas, R., Madjovski, A., Ruhrmann, S., Faigle, W., Frauenknecht, K., Pinilla, C., Santos, R., Hammer, C., Ortiz, Y., Opitz, L., Gronlund, H., Rogler, G., Boyman, O., Reynolds, R., Lutterotti, A., Khademi, M., Olsson, T., Piehl, F., Sospedra, M. & Martin, R. (2018) Memory B Cells Activate Brain-Homing, Autoreactive CD4(+) T Cells in Multiple Sclerosis. Cell, 175(1), 85-100.e23.

Kappos, L., Li, D., Calabresi, P. A., O’Connor, P., Bar-Or, A., Barkhof, F., Yin, M., Leppert, D., Glanzman, R., Tinbergen, J. & Hauser, S. L. (2011) Ocrelizumab in relapsing-remitting multiple sclerosis: a phase 2, randomised, placebo-controlled, multicentre trial. Lancet, 378(9805), 1779–87.

Marjolein, W., Lisman, J., Hoebert, J., Moltó Puigmarti, C., Dijk, L. v., Langedijk, J., Marchange, S., Damen, N., Vervloet, M. & Directorate-General for Health and Food Safety (European Commission) (2019) Study on off-label use of medicinal products in the European Union : report.

Memon, A. B., Javed, A., Caon, C., Srivastawa, S., Bao, F., Bernitsas, E., Chorostecki, J., Tselis, A., Seraji-Bozorgzad, N. & Khan, O. (2018) Long-term safety of rituximab induced peripheral B-cell depletion in autoimmune neurological diseases. PLoS One, 13(1), e0190425.

Montalban, X., Hauser, S. L., Kappos, L., Arnold, D. L., Bar-Or, A., Comi, G., de Seze, J., Giovannoni, G., Hartung, H.-P., Hemmer, B., Lublin, F., Rammohan, K. W., Selmaj, K., Traboulsee, A., Sauter, A., Masterman, D., Fontoura, P., Belachew, S., Garren, H., Mairon, N., Chin, P. & Wolinsky, J. S. (2017) Ocrelizumab versus Placebo in Primary Progressive Multiple Sclerosis. The New England Journal of Medicine.

Nielsen, A. S., Miravalle, A., Langer-Gould, A., Cooper, J., Edwards, K. R. & Kinkel, R. P. (2012) Maximally tolerated versus minimally effective dose: the case of rituximab in multiple sclerosis, Mult Scler. England, 377–8.

Palanichamy, A., Jahn, S., Nickles, D., Derstine, M., Abounasr, A., Hauser, S. L., Baranzini, S. E., Leppert, D. & von Budingen, H. C. (2014) Rituximab efficiently depletes increased CD20-expressing T cells in multiple sclerosis patients. J Immunol, 193(2), 580–586.

Rissanen, E., Remes, K. & Airas, L. (2017) Severe neutropenia after rituximab-treatment of multiple sclerosis. Mult Scler Relat Disord, 20, 3–5.

Rommer, P. S., Dorner, T., Freivogel, K., Haas, J., Kieseier, B. C., Kumpfel, T., Paul, F., Proft, F., Schulze-Koops, H., Schmidt, E., Wiendl, H., Ziemann, U. & Zettl, U. K. (2016) Safety and Clinical Outcomes of Rituximab Treatment in Patients with Multiple Sclerosis and Neuromyelitis Optica: Experience from a National Online Registry (GRAID). J Neuroimmune Pharmacol, 11(1), 1–8.

Salzer, J., Lycke, J., Wickstrom, R., Naver, H., Piehl, F. & Svenningsson, A. (2016a) Rituximab in paediatric onset multiple sclerosis: a case series. J Neurol, 263(2), 322–326.

Salzer, J., Svenningsson, R., Alping, P., Novakova, L., Björck, A., Fink, K., Islam-Jakobsson, P., Malmeström, C., Axelsson, M., Vågberg, M., Sundström, P., Lycke, J., Piehl, F. & Svenningsson, A. (2016b) Rituximab in multiple sclerosis: A retrospective observational study on safety and efficacy. Neurology, 87(20), 2074–2081.

Scotti, B., Disanto, G., Sacco, R., Guigli, M., Zecca, C. & Gobbi, C. (2018) Effectiveness and safety of Rituximab in multiple sclerosis: an observational study from Southern Switzerland. PLoS One, 13(5), e0197415.

Sorensen, P. S., Lisby, S., Grove, R., Derosier, F., Shackelford, S., Havrdova, E., Drulovic, J. & Filippi, M. (2014) Safety and efficacy of ofatumumab in relapsing-remitting multiple sclerosis: a phase 2 study. Neurology, 82(7), 573–81.

Svenningsson, A., Bergman, J., Dring, A., Vagberg, M., Birgander, R., Lindqvist, T., Gilthorpe, J. & Bergenheim, T. (2015) Rapid depletion of B lymphocytes by ultra-low-dose rituximab delivered intrathecally. Neurol Neuroimmunol Neuroinflamm, 2(2), e79.

Topping, J., Dobson, R., Lapin, S., Maslyanskiy, A., Kropshofer, H., Leppert, D., Giovannoni, G. & Evdoshenko, E. (2016) The effects of intrathecal rituximab on biomarkers in multiple sclerosis. Mult Scler Relat Disord, 6, 49–53.

Working group appointed by the Finnish Medical Society Duodecim and the Finnish Neurological Society (2019) Finnish Current Care Guideline Multiple sclerosis, 2019. Available online: https://www.kaypahoito.fi/ [Accessed 18 Nov 2019]

